# Booster dose of self-amplifying SARS-CoV-2 RNA vaccine vs. mRNA vaccine: a phase 3 comparison of ARCT-154 with Comirnaty^®^

**DOI:** 10.1101/2023.07.13.23292597

**Authors:** Yoshiaki Oda, Yuji Kumagai, Manabu Kanai, Yasuhiro Iwama, Iori Okura, Takeshi Minamida, Yukihiro Yagi, Toru Kurosawa, Benjamin Greener, Ye Zhang, Judd L. Walson

**Affiliations:** Meiji Seika Pharma Co., Ltd., Chuo-ku, Tokyo, Japan; Kitasato University Kitasato Institute Hospital, Minato-ku, Tokyo, Japan; Arcturus Therapeutics, Inc., San Diego, CA, USA

## Abstract

**Background:** Licensed mRNA vaccines demonstrated initial effectiveness against COVID-19 but require booster doses to broaden the anti-SARS-CoV-2 response. There is an unmet need for novel highly immunogenic and broadly protective vaccines. We compared immunogenicity and tolerability of ARCT-154, a novel self-amplifying mRNA vaccine with the mRNA vaccine, Comirnaty^®^.

**Methods:** We compared immune responses to ARCT-154 and Comirnaty booster doses in healthy 18– 77-year-old Japanese adults initially immunised with two doses of mRNA COVID-19 vaccine (Comirnaty or Spikevax^®^) then a third dose of Comirnaty at least 3 months previously. Neutralising antibodies were measured before and 28 days after booster vaccination. The primary objective was to demonstrate non-inferiority of the immune response against Wuhan-Hu-1 SARS-CoV-2 virus as geometric mean titre (GMT) ratios and seroresponse rates (SRR) of neutralising antibodies; key secondary endpoints included the immune response against the Omicron BA.4/5 variant and vaccine tolerability assessed using participant-completed electronic diaries.

**Findings:** Between December 13, 2022 and February 25, 2023 we enrolled 828 participants randomised 1:1 to receive ARCT-154 (n = 420) or Comirnaty (n = 408) booster doses. Four weeks after boosting, ARCT-154 induced higher Wuhan-Hu-1 neutralising antibodies GMTs than Comirnaty (5641 [95% CI: 4321, 7363] and 3934 [2993, 5169], respectively), a GMT ratio of 1·43 (95% CI: 1·26–1·63), with SRR of 65·2% (60·2–69·9) and 51·6% (46·4–56·8) meeting the non-inferiority criteria. Respective anti-Omicron BA.4/5 GMTs were 2551 (1687–3859) and 1958 (1281–2993), a GMT ratio of 1·30 (95% CI: 1·07–1·58), with SRR of 69·9% (65·0–74·4) and 58·0% (52·8–63·1), meeting the superiority criteria for ARCT-154 over Comirnaty. Booster doses of either ARCT-154 or Comirnaty were equally well-tolerated with no causally-associated severe or serious adverse events; 94·8% and 96·8% of ARCT-154 and Comirnaty vaccinees reported local reactions and 65·7% and 62·5% had solicited systemic adverse events. Events were mainly mild in severity, occurring and resolving within 3–4 days of vaccination.

**Interpretation:** Immune responses four weeks after an ARCT-154 booster dose in mRNA-immunised adults were higher than after a Comirnaty booster, meeting non-inferiority criteria against the prototype Wuhan-Hu-1 virus, and superiority criteria against the Omicron BA.4/5 variant.

**Funding:** The study was funded by the Japanese Ministry of Health, Labour, and Welfare following a public invitation to bid for an urgent improvement project for vaccine manufacturing systems, fourth invitation, Grant number: 1212-3.

**Clinical Trials registration and identifier:** The study was registered on the Japan Registry for Clinical Trials (jRCT 2071220080).

## INTRODUCTION

Although global rates of infections caused by the severe acute respiratory syndrome coronavirus 2 (SARS-CoV-2) have decreased since the peak incidences of disease in early 2022, millions of cases continue to occur, primarily driven by the emergence of new variants [1]. Many effective vaccines have been developed to protect against the more severe consequences of SARS-CoV-2 but they are proving to be less effective against the new variants which increasingly predominate [2]. This is a consequence of the combined effects of waning antibody titres following primary immunisation and the lower sensitivity of the newly emergent variants and sub-variants to antibodies elicited against the virus from which the first vaccines were developed [3]. Lower sensitivity is a result of the successive accumulation of mutations in the spike protein (S-protein), the main antigenic target of COVID-19 vaccines [4], altering the epitopes on the glycoprotein to make them less susceptible to vaccine-induced neutralising antibodies. Strategies to overcome these factors and maintain immune protection include administration of booster doses, development of new vaccine formulations based on more recent strains, and use of heterologous immunisation with different vaccines to broaden and prolong the immune response.

Some of the most widely used vaccines are based on mRNA coding for the S-protein which have been proven to be effective, but also to have a limited duration of immune response [5,6]. Homologous boosting of mRNA COVID-19 vaccines extends the response but does not broaden the response against antigenic drift that occurs as novel variants emerge [7–9]. New booster formulations incorporating antigen or mRNA directly targeting the new variants have been introduced [10]. However, novel vaccines may be necessary to extend protective immunity initially established by the first vaccines and cover as yet unknown new variants. Arcturus (Arcturus Therapeutics Inc., San Diego, CA, USA) has developed an alternative technology for mRNA vaccination using self-amplifying RNA vaccines (sa-mRNA). sa-mRNA vaccines appear to require lower doses for enhanced antigen expression compared with conventional mRNA-based vaccines and may improve the efficacy and duration of protection [11–13]. One such formulation, ARCT-154, encodes the S protein of B.1 with the D614G mutation, one of the earliest detected variants of SARS-CoV-2 [14]. In collaboration with Meiji Seika Pharma Co., Ltd. (Tokyo, Japan), who are co-developing and marketing the vaccine in Japan, we have assessed the safety, tolerability and immunogenicity of ARCT-154 in comparison with the mRNA vaccine Comirnaty^®^ (Pfizer Co. Ltd./BioNTech) when used as a booster dose in healthy adults who have previously been immunised with three doses of approved mRNA vaccines and Comirnaty as their last (third) COVID-19 vaccination. This study follows the primary phase 1/2/3a/3b study (ClinicalTrials.gov: NCT05012943), which evaluated the safety and efficacy of ARCT-154 (manuscript in preparation).

## METHODS

This randomised, phase 3, double-blind, active-controlled comparative study was conducted in 11 sites in Japan (*see supplementary table 1*) from December 2022 to February 2023. The study protocol was approved by the institutional review boards of all participating sites and was registered on the Japan Registry for Clinical Trials (jRCT 2071220080). The study was done according to the ethical principles based on the Declaration of Helsinki and Good Clinical Practice. All participants provided written informed consent before enrolment. The primary objective was to demonstrate the non-inferiority of the immune response against SARS-CoV-2 (Wuhan strain) four weeks after an ARCT-154 booster dose compared with that elicited by a Comirnaty booster in mRNA-immunised adults using two endpoints, the geometric mean titre (GMT) of neutralising antibodies and the seroresponse rate (SRR). A similar assessment of the non-inferiority of the immune response against Omicron BA.4/5 using the same endpoints, and the safety and tolerability of vaccination were key secondary objectives.

### Participants

Eligible participants were healthy adults 18 years of age or older who had previously been immunised with three documented doses of one of two authorized mRNA vaccines, either Comirnaty or Spikevax^®^ (Moderna Inc.), with the last dose being Comirnaty received at least 3 months previously. All enrolled participants had to agree to comply with study requirements including all study visits and provision of blood samples, and to adhere to contraceptive requirements from 28 days before study vaccination and then throughout the study duration.

The main exclusion criteria were any indication of a current infection, e.g., temperature ≥ 37·5°C on Day 1 or any known history of a COVID-19 infection within the previous 4 months, any chronic infection such as HIV, HBV, HCV, or active tuberculosis or any history of myocarditis, pericarditis, cardiomyopathy, or medical or psychological issue that, based on the medical judgement of the principal investigator, put the volunteer at risk or risked preventing completion of the study. Other exclusion criteria included any known immunosuppressive condition or treatment likely to influence immune responses to vaccination, any known history of adverse reactions to vaccination, any participation in any other drug or vaccine trial, or a positive pregnancy test at the time of screening or intention to become pregnant within one year of vaccination.

### Vaccines

The ARCT-154 study vaccine consists of sa-mRNA encapsulated in lipid nanoparticles. The RNA comprises a replicon based upon Venezuela equine encephalitis virus (VEEV) in which RNA coding for the VEEV structural proteins has been replaced with RNA coding for the full-length spike (S) glycoprotein of the SARS-CoV-2 D614G variant. Vaccine was supplied in a vial containing 100 μg active ingredient stored at -20°C or lower before use, which was dissolved in 10 mL sterile saline and a 0·5 mL dose containing 5 μg administered by intramuscular injection in the deltoid.

The control mRNA vaccine, Comirnaty was supplied as a frozen suspension in vials containing 225 μg of a nucleoside-modified mRNA encoding the viral spike (S) glycoprotein of SARS-CoV-2 in 0.45 mL sterile solution stored at -80°C. After further diluting each vial with 1·8 mL saline the recommended 0·3 mL booster dose of Comirnaty containing 30 μg was administered by intramuscular injection in the deltoid. Vaccines were prepared and administered by study staff who were unblinded to the randomisation code supplied by the study sponsor and who played no further role in the study. Collection of blood samples and safety data was done by blinded study staff and all subsequent procedures, including immunogenicity analyses, were done in a blinded manner. Participants were also unaware of their vaccine allocation.

### Procedures

On Day 1, enrolled participants had a medical examination and a baseline blood draw, before receiving their assigned vaccine. After monitoring for 30 minutes for any immediate reactions, participants recorded solicited reactogenicity in electronic diaries for 7 days. Solicited local reactions (injection site pain, tenderness, swelling, erythema and induration) and systemic adverse events (pyrexia, arthralgia, chills, diarrhoea, dizziness, headache, malaise, nausea, vomiting, myalgia) were assessed for severity (*see supplementary table 2*). Participants continued to record the occurrence of any unsolicited adverse events and symptoms of interest which included chest pain or shortness of breath to monitor for potential symptoms that could suggest myocarditis and pericarditis. Any serious adverse event (SAE) were reported immediately to the investigator. Electronic diaries were collected at the second study visit on Day 29, when study staff reviewed them to ensure they had been completed correctly and the investigator assessed the causal relationship of any adverse events with vaccination. Safety monitoring up to 12 months is ongoing in this study; only adverse events reported up to Day 29 are included in this interim report.

### Immunogenicity analyses

Sera were prepared immediately from blood samples drawn on Days 1 and 29 and stored at - 20°C for shipping to the Labcorp Central Laboratory Services LP (Indianapolis, IN, USA) for measurement of neutralising antibodies against Wuhan-Hu-1 and Omicron BA.4/5 sublineage SARS-CoV-2 pseudoviruses. Antibodies were expressed as group GMTs at Days 1 and 29, for which initially seronegative samples were assigned a value of half the lower limit of quantitation (LLOQ) which was a dilution of 1:40. SRR were calculated at Day 29, being the group proportions demonstrating either a fourfold or greater increase in titre from Day 1 to Day 29 in initially seropositive participants, or a four-fold higher titre than half the LLOQ for initially seronegative participants. Sera were also used for a qualitative determination of antibodies against the SARS-CoV-2 nucleocapsid protein using a commercial test kit (Cica Immunotest SARS-CoV-2 IgG EX, Kanto Chemical Co., Inc.); a positive result was considered to be an indicator of a prior SARS-CoV-2 infection.

### Statistics

#### Analysis subsets

Primary analyses were performed in a per protocol subset, PPS-1, and sensitivity analyses were done in a second subset, PPS-2. Both subsets consisted of participants with no protocol violations related to eligibility criteria, dosage and administration, concomitant drugs/therapies, immunogenicity data, and confirmation items on the day of study vaccine administration. PPS-1 consists of those who were seronegative for the nucleocapsid of SARS-CoV-2 before study vaccine administration, which is considered to indicate no recent prior infection, PPS-2 includes both those seropositive and seronegative for SARS-CoV-2 nucleocapsid. Descriptive analyses of safety endpoints were determined in the Safety Set consisting of all participants who received a study vaccination and who had safety data and no significant GCP deviations.

#### Sample size

For the immunogenicity evaluations, the GMT ratio of the neutralising antibody titre to the conventional strain in the booster vaccination was assumed to be 1·0 with a standard deviation of 0·40, and the SRRs of each group was assumed to be 85%. To obtain a statistical power of 90% to detect non-inferiority of ARCT-154 to Comirnaty in GMT and SRR (GMT non-inferiority margin = 0·67, SRR non-inferiority margin = -10%, significance level = 1- sided 2·5%), 270 participants per group were required for analysis. Assuming a 10% drop-out rate from immunogenicity analyses would require a total of 600 participants, and assuming a further 20% of randomised participants being excluded for being seropositive to the SARS-CoV-2 nucleocapsid before study vaccine administration a total of 780 participants were needed.

#### Immunogenicity analyses

The primary objective was to demonstrate the non-inferiority of the immune response to ARCT-154 when compared with Comirnaty as both the neutralising antibody GMT and SRR against SARS-CoV-2 (Wuhan-Hu-1 strain) 28 days after vaccination in the per protocol PPS-1 subset. For GMTs, analysis of covariance (ANCOVA) was conducted for the log-transformed neutralising titres against Wuhan-Hu-1 strain SARS-CoV-2 on Day 29 with the group as a factor, the allocation factor (time since the last vaccination as a categorical variable [less than 5 months, 5 months and beyond], gender, age as a continuous variable) as covariates. The geometric mean ratio (GMR) of neutralising antibody titres against SARS-CoV-2 at Day 29 in ARCT-154 compared with Comirnaty was calculated with its 95% CI by inverting LS-means difference between ARCT-154 and Comirnaty using this ANCOVA model; non-inferiority was confirmed when the lower bound of the inverted 95% CI exceeded 0.67.

The difference in SRR for SARS-CoV-2 (Wuhan-Hu-1 strain) on Day 29 between ARCT-154 and Comirnaty and its 95% CI were calculated by the Miettinen-Nurminen method, with a randomisation factor (time since the last vaccination as a categorical variable [less than 5 months, 5 months and beyond], gender, age as a continuous variable) as an adjustment factor; non-inferiority of ARCT-154 to Comirnaty was confirmed if the lower limit of the 95% CI exceeded -10%.

#### Analysis of Immunogenicity Secondary Endpoints

Major secondary endpoints included the GMT and SRR of neutralising antibodies against SARS-CoV-2 (Omicron BA.4/5 strain) pseudovirus on Day 29, including a non-inferiority evaluation of ARCT-154 to Comirnaty on Day 29. If non-inferiority was shown, then superiority of ARCT-154 to Comirnaty was evaluated; superiority was confirmed if the anti-log transformed GMT ratio lower bound of the 95% CI was greater than 1. We also calculated geometric mean-fold rises (GMFR) in neutralising antibody titres against both Wuhan-Hu-1 and Omicron BA.4/5 strains of SARS-CoV-2 on Day 29 relative to Day 1 before vaccine administration, and summary statistics of fold rise with 95% CIs are presented for each group.

#### Safety assessments

Incidences of solicited adverse events (AEs), symptoms of interest and any additional symptoms occurring after administration up to Day 7, and unsolicited AEs, deaths, serious AEs (SAEs), medically significant AEs, and AEs of special interest up to Day 29 were calculated in the Safety Set and presented descriptively with causal relationship with study vaccine as assessed by the investigator.

### Role of the Funder

The funder of the study had no role in the study design, data collection, data analysis, data interpretation, or writing of the report.

## RESULTS

A total of 986 volunteers were screened, of whom 828 were enrolled into the study. The 158 screening failures were mainly due to high blood pressure or COVID-19 positivity. Enrolled volunteers were randomly allocated to the ACRT-154 (n = 420) and Comirnaty (n = 408) groups. All enrolees who received their vaccination according to the randomisation were included the in the Safety Set (**Table 1**). Mean age (± SD) was 45·7 (± 11·8) years, ranging from 18 to 77 years, and there were more female than male participants (485 vs. 340). The overwhelming majority (810/825 [98·2%]) had received their last COVID-19 vaccination at least 5 months earlier but 817 of 825 (99·0%) still had neutralising antibodies against Wuhan-Hu-1 SARS-CoV-2, while 654 of 825 (79·3%) had neutralising antibodies against Omicron BA.4/5 prevaccination. Relatively few participants, 56 of 818 (6·8%) were seropositive for SARS-CoV-2 nucleocapsid protein, which is an indicator of infection by the virus. All participants had received Comirnaty as their last (third) COVID-19 vaccination. Most participants (79·8% [658/825]) had a history of exclusive Comirnaty vaccination and 20·1 % (166/825) had received two doses of Spikevax before the third booster Comirnaty dose.

**Table 1.** Demographics of the study population in the Full Analysis Set.

| Parameter |  | ARCT-154 | Comirnaty | Total |
| --- | --- | --- | --- | --- |
|  |  | N = 417 | N = 408 | N = 825 |
| **Age, years** | Mean (± SD) | **45·2** (12·0) | **46·2** (11·6) | **45·7** (11·8) |
|  | [Range] | [18·0, 77·0] | [18·0, 76·0] | [18·0, 77·0] |
| < 65 years | n (%) | 405 (97·1) | 400 (98·0) | 805 (97·6) |
| ≥ 65 years | n (%) | 12 (2·9) | 8 (2·0) | 20 (2·4) |
| **Gender, n (%)** | Female | 246 (59·0) | 239 (58·6) | 485 (58·8) |
|  | Male | 171 (41·0) | 169 (41·4) | 340 (41·2) |
| **Time since 3rd vaccination, n (%)** | < 5 months | 11 (2·6) | 4 (1·0) | 15 (1·8) |
| | ≥ 5 months | 406 (97·4) | 404 (99·0) | 810 (98·2) |
| **Participants requiring caution in vaccination, n (%)** | Underlying disease | 72 (17·3) | 62 (15·2) | 134 (16·2) |
|  | Previous symptoms indicative of allergic reaction # | 90 (21·6) | 88 (21·6) | 178 (21·6) |
|  | History of convulsions | 6 (1·4) | 1 (0·2) | 7 (0·8) |
| **Neutralising antibodies at baseline** |  |  |  |  |
| **Wuhan-Hu-1, n (%)** | Seronegative | 5 (1·2) | 3 (0·7) | 8 (1·0) |
|  | Seropositive | 412 (98·8) | 405 (99·3) | 817 (99·0) |
| **Omicron BA.4/5, n (%)** | Seronegative | 84 (20·1) | 87 (21·3) | 171 (20·7) |
|  | Seropositive | 333 (79·9) | 321 (78·7) | 654 (79·3) |
| **SARS-CoV-2 nucleocapsid antibody, n (%)** | Seronegative | 388 (93·0) | 381 (93·4) | 769 (93·2) |
|  | Seropositive | 29 (7·0) | 27 (6·6) | 56 (6·8) |
| **Previous brands of vaccine received, n (%)**<br>* | C + C + C | 329 (78·9) | 329 (80·6) | 658 (79·8) |
|  | C + S + C | 0 (0) | 0 (0) | 0 (0) |
|  | S + C + C | 1 (0·2) | 0 (0) | 1 (0·1) |
|  | S + S + C | 87 (20·9) | 79 (19·4) | 166 (20·1) |

### Immunogenicity

Background immunity against Wuhan-Hu-1 SARS-CoV-2 was evident in both groups, with similar baseline GMTs in both groups in the PPS-1 (**Table 2**). Four weeks after the study booster doses, both groups displayed marked increases in neutralising antibody titres, from 813 (95% CI: 716, 924) to 5641 (4321, 7363) after ARCT-154 and from 866 (755, 993) to 3934 (2993, 5169) after Comirnaty, representing geometric mean-fold rises of 6·7 and 4·4, respectively. The first primary immunogenicity endpoint, the GMT ratio at Day 29 of neutralising antibodies after ARCT-154 compared with Comirnaty, was 1·43 (95% CI: 1·26, 1·63). The lower limit of the 95% CI exceeded the predefined non-inferiority margin of 0·67, confirming non-inferiority of the ARCT-154 response (**Table 2**). Similar analyses of immune response against Wuhan-Hu-1 in the FAS and the PPS-2 subsets also showed non-inferiority of the ARCT-154 response (*see supplementary tables 4 and 5*).

**Table 2.**
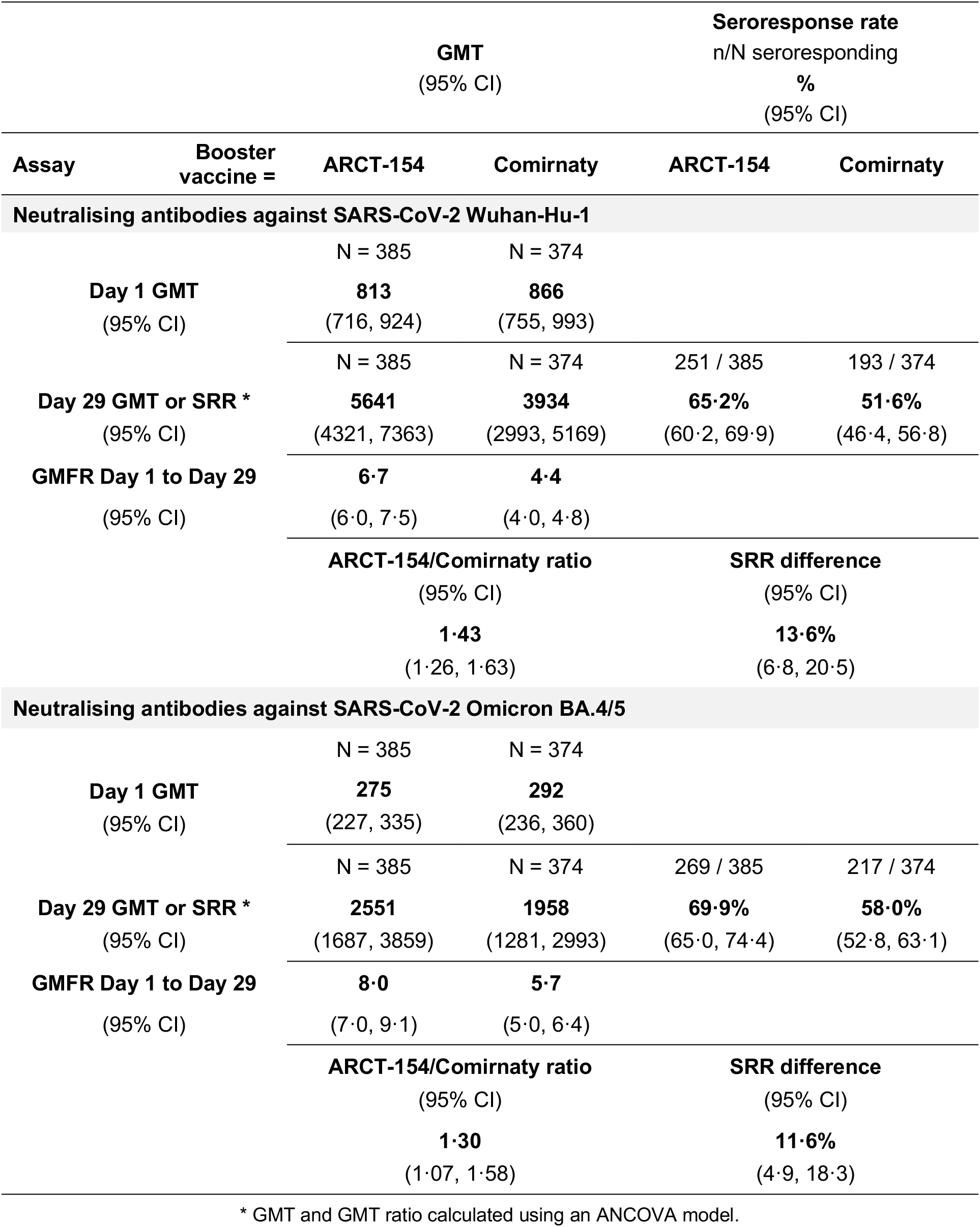
Geometric mean titres (GMT) of neutralising antibodies at baseline on Day 1, and GMTs and seroresponse rates (SRR) at Day 29 with non-inferiority comparisons after ARCT-154 and Comirnaty (PPS-1)

| Assay | Booster vaccine = | GMT<br>(95% CI) |  | Seroresponse rate<br>n/N seroresponding<br>%<br>(95% CI) |  |
| --- | --- | --- | --- | --- | --- |
|  |  | ARCT-154 | Comirnaty | ARCT-154 | Comirnaty |
| Neutralising antibodies against SARS-CoV-2 Wuhan-Hu-1 |  |  |  |  |  |
|  |  | N = 385 | N = 374 |  |  |
| Day 1 GMT<br>(95% CI) |  | 813<br>(716, 924) | 866<br>(755, 993) |  |  |
|  |  | N = 385 | N = 374 | 251 / 385 | 193 / 374 |
| Day 29 GMT or SRR *<br>(95% CI) |  | 5641<br>(4321, 7363) | 3934<br>(2993, 5169) | 65.2%<br>(60.2, 69.9) | 51.6%<br>(46.4, 56.8) |
| GMFR Day 1 to Day 29<br>(95% CI) |  | 6.7<br>(6.0, 7.5) | 4.4<br>(4.0, 4.8) |  |  |
|  |  | ARCT-154/Comirnaty ratio<br>(95% CI) |  | SRR difference<br>(95% CI) |  |
|  |  | 1.43<br>(1.26, 1.63) |  | 13.6%<br>(6.8, 20.5) |  |
| Neutralising antibodies against SARS-CoV-2 Omicron BA.4/5 |  |  |  |  |  |
|  |  | N = 385 | N = 374 |  |  |
| Day 1 GMT<br>(95% CI) |  | 275<br>(227, 335) | 292<br>(236, 360) |  |  |
|  |  | N = 385 | N = 374 | 269 / 385 | 217 / 374 |
| Day 29 GMT or SRR *<br>(95% CI) |  | 2551<br>(1687, 3859) | 1958<br>(1281, 2993) | 69.9%<br>(65.0, 74.4) | 58.0%<br>(52.8, 63.1) |
| GMFR Day 1 to Day 29<br>(95% CI) |  | 8.0<br>(7.0, 9.1) | 5.7<br>(5.0, 6.4) |  |  |
|  |  | ARCT-154/Comirnaty ratio<br>(95% CI) |  | SRR difference<br>(95% CI) |  |
|  |  | 1.30<br>(1.07, 1.58) |  | 11.6%<br>(4.9, 18.3) |  |

\* GMT and GMT ratio calculated using an ANCOVA model.

For the seroresponse rate endpoint on Day 29, the difference in SRR for neutralising antibodies against SARS-CoV-2 (Wuhan-Hu-1 strain) of ARCT-154 and Comirnaty was 13·6% (95% CI: 6·8, 20·5). Since the lower limit of the 95% CI exceeded the predefined non-inferiority margin of -10%, the primary objective of non-inferiority of the immune response to ARCT-154 as booster dose compared with Comirnaty by both assessment measures, GMT and SRR, was confirmed.

In addition, the secondary GMT and SRR endpoints of the response against the Omicron BA.4/5 variant also demonstrated non-inferiority of the ARCT-154 response compared with the Comirnaty response (**Table 2**). Respective GMTs of 275 (95% CI: 227, 335) and 292 (2360, 360) against Omicron BA.4/5 at Day 1 in ARCT-154 and Comirnaty groups were increased to 2551 (1687, 3859) and 1958 (1281, 2993) by Day 29 by Day 29, with respective GMFRs of 8·0 and 5·7. The GMT ratio for ARCT-154 to Comirnaty groups was 1·30 (95% CI: 1·07, 1·58), the lower limit of the 95% CI exceeding the non-inferiority criterion of 0·67, and the difference in SRR was 11·6% (4·9, 18·3) exceeding the -10% non-inferiority. Importantly, as the lower limit of the 95% CI for the GMT ratio was greater than 1.0, and the lower limit of the 95% CI for the SRR difference was greater than 0, which met the criteria for superiority of the ARCT-154 response over the Comirnaty response.

The response was not affected by the previous mRNA vaccination history, i.e., whether the three required mRNA vaccinations were all Comirnaty or some were Spikevax. In participants previously immunised with three doses of Comirnaty the GMT after ARCT-154 was 5124 (95% CI: 3735, 7031; n = 306) and 3391 (2443, 4707; n = 298) after a fourth dose of Comirnaty, a GMT ratio of 1·51 (1·31, 1·74). After previously receiving two doses of Spikevax and one of Comirnaty, the GMT after ARCT-154 was 7040 (95% CI: 4270, 11608; n = 78) and 5751 (3534, 9357; n = 76) after a booster dose of Comirnaty, a GMT ratio of 1·22 (0·94, 1·59).

Non-inferiority of the ARCT-154 response compared with Comirnaty was also consistently observed when groups were further separated according to age category (< 65 years vs. ≥ 65 years), gender (male vs. female), or interval since the last booster COVID-19 vaccination before the study vaccination was administered (< 5 months vs. ≥ 5 months) (*Supplementary tables 4-6*).

### Safety and tolerability

Both vaccines were well tolerated as booster doses. Up to the interim cut-off of Day 29 presented in this analysis, there were no deaths, and no adverse events of special interest or medically-attended adverse events reported (**Table 3**). One serious adverse event (SAE) reported in a participant in the Comirnaty group was described as a foot deformity and was not considered to have any causal relationship to the study vaccination by the investigators. There were no reports of solicited local reactions or systemic adverse events described as grade 4 or life threatening. Solicited local reactions were reported by 94·8% (398/420) of the ARCT-154 vaccinees and 96·8% (395/408) of those who received Comirnaty in the Safety Set (**Table 3**). Most reported local reactions were transient, starting within 1 or 2 days of vaccination and resolving by day 4 (*see supplementary table 7*), and were described as mild or moderate in severity. The majority in both groups were mild pain and/or tenderness at the injection site (**Figure 2**). Seven participants reported severe local reactions, 3 participants reported 4 severe local reactions after ARCT-154 – individual cases of severe pain, tenderness, swelling and induration at the injection site. Four Comirnaty recipients reported 5 severe local reactions, including 3 cases of severe erythema, swelling and tenderness at the injection site. Swelling, erythema and induration were reported less frequently than pain or tenderness, but all three reactions were more frequent (23·8%, 20·8% and 19·9%) after Comirnaty than ARCT-154 (14·0%, 12·4% and 12·4%).

**Figure 1.**
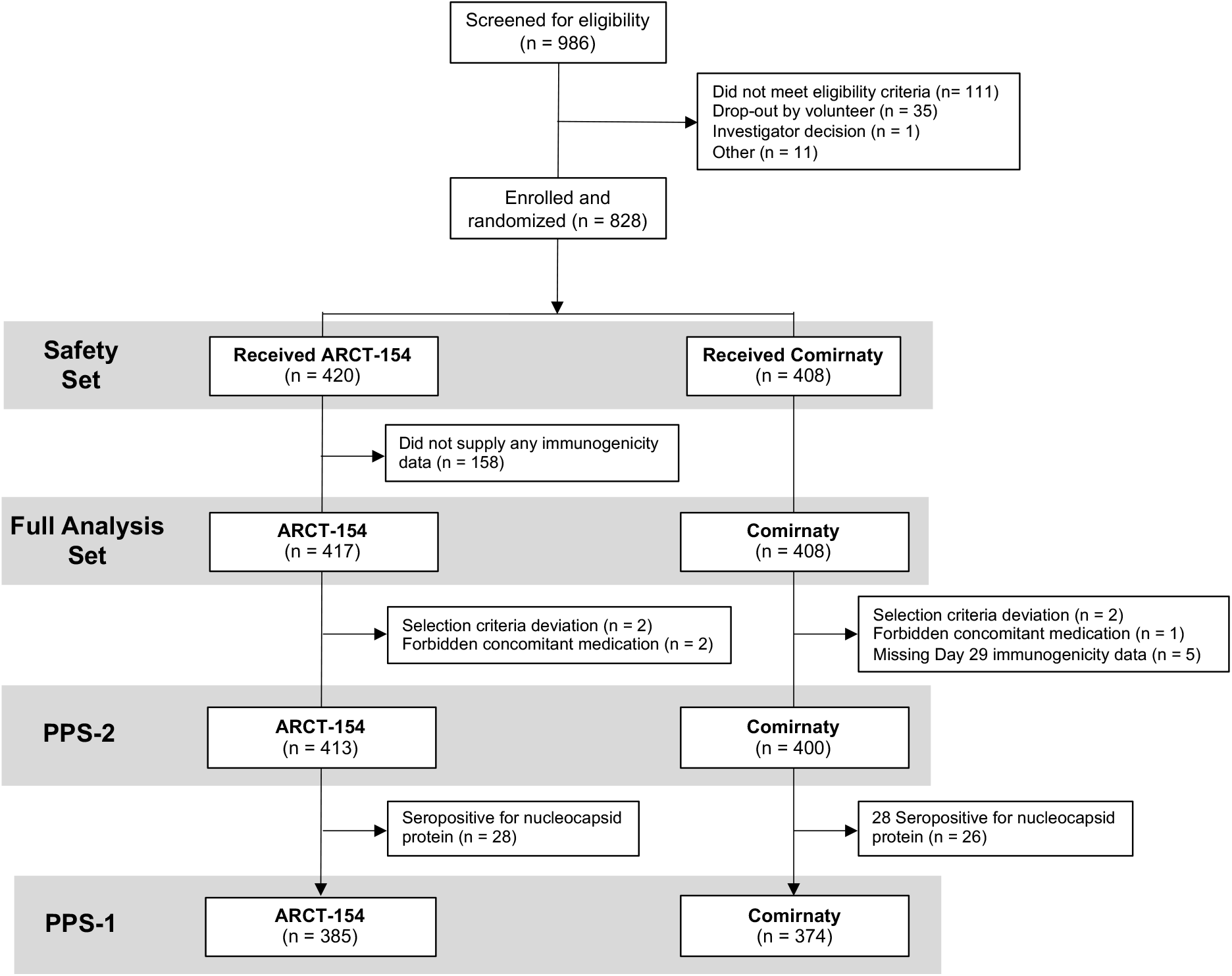
Study flow chart showing eligibility for analysis groups.

**Figure 2.**
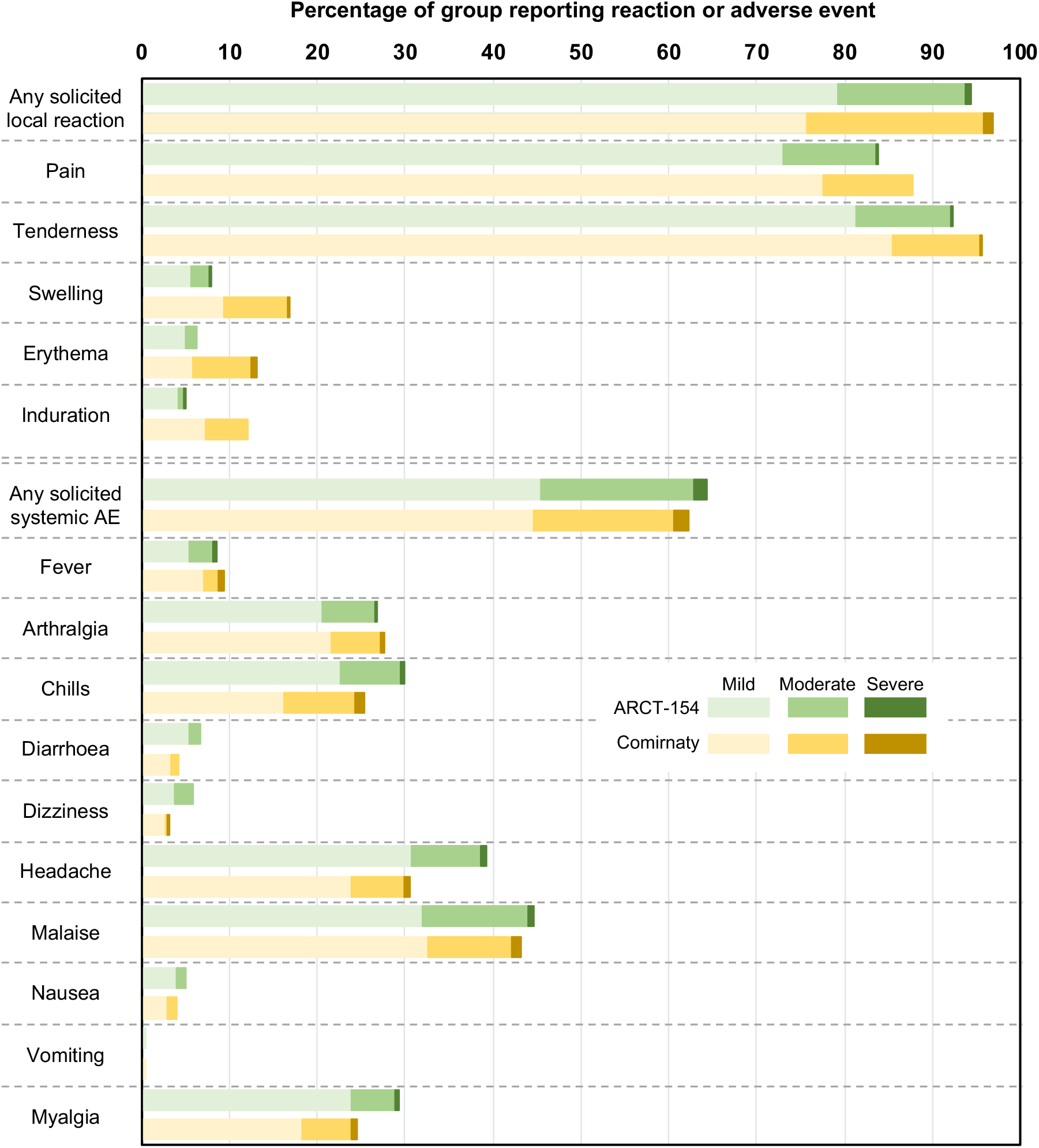
Rates of solicited local reactions and systemic adverse events in the two study groups with severity.

**Table 3.**
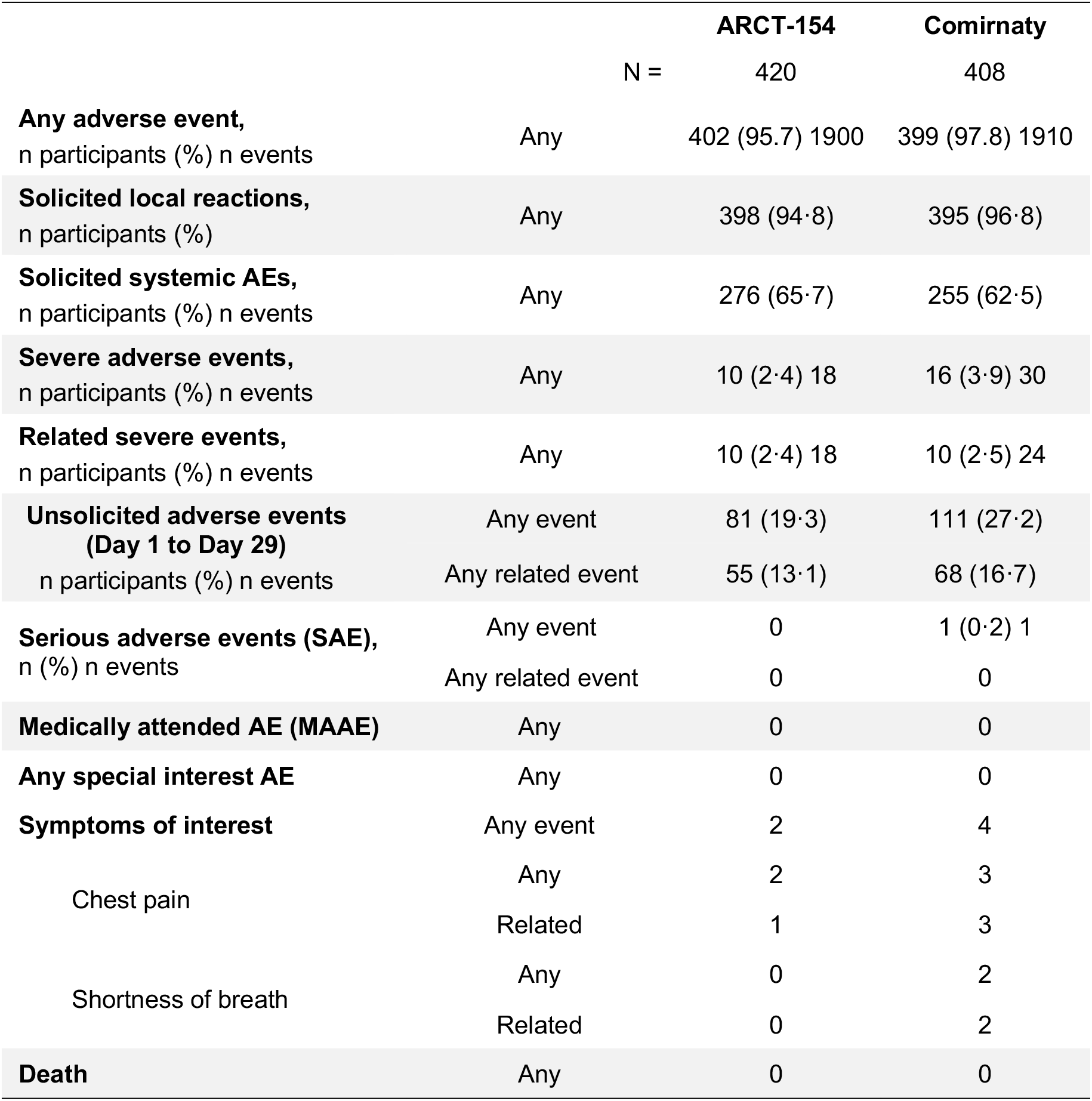
Rates of adverse events (AE), serious adverse events (SAE), adverse events of special interest and MAAE in the two study groups from Day 1 to Day 29 (Safety Set)

Solicited systemic adverse events were reported by 65·7% (276/420) and 62·5% (255/408) of ARCT-154 and Comirnaty vaccinees, respectively (**Table 3**). The most frequent was malaise, reported by 44·8% (188 of 420) and 43·1% (176 of 408) of ARCT-154 and Comirnaty groups, followed by headache reported by 39·3% (165 of 420) and 30·6% (125 of 408), chills by 30·0% (126 of 420) and 25·2% (103 of 408), myalgia by 29·3% (123 of 420) and 24·5% (100 of 408), and arthralgia reported by 26·7% (112 of 420) and 27·7% (113 of 408). As with the local reactions, solicited systemic adverse events were mainly mild and transient (**Figure 2**), starting and resolved within 1 to 3 days of vaccination (*see supplementary table 8*).

Unsolicited adverse events during and the postvaccination period up to Day 29 were reported by 19·3% (81 of 420) of ARCT-154 recipients and 27·2% (111 of 408) of the Comirnaty group (**Table 3**). Of these the unsolicited AEs in 13·1% (55 of 420) and 16·7% (68 of 408) of ARCT-154 and Comirnaty groups were considered to be causally related to the vaccinations by the investigators. Frequencies of disorders listed according to the MedDRA classifications are shown in *supplementary table 9*. The majority of the reported unsolicited AEs were mild or moderate but there were seven AEs described as severe (Grade 3), one in the ARCT-154 group and six in the Comirnaty group. The severe unsolicited AE in the ARCT-154 group was a case of abnormal hepatic function which was assessed as related to the investigative vaccine by the investigators. The six severe unsolicited AEs in Comirnaty recipients were three cases of nasopharyngitis, and individual cases of pyrexia, ankle fracture and foot deformity, none of which was considered to be related to the vaccine.

Occurrences of the symptoms of interest, chest pain and shortness of breath, between days 1 and 7 after vaccination were specifically monitored as indicators of potential myocarditis and pericarditis. Two ARCT-154 recipients reported chest pain (**Table 3**); one within 1 day of vaccination which was the investigator considered related to the vaccine, and the second 6 days after vaccination which was considered unrelated. Four of the Comirnaty recipients reported five symptoms – three cases of chest pain occurring within 1–3 days of vaccination and two cases of shortness of breath occurring within a day of vaccination—all of which were assessed as related to the study vaccination by investigators. With follow up, no indication of myocarditis or pericarditits was detected in participants in either vaccine group.

## DISCUSSION

In this analysis of an ongoing study in healthy adults previously immunised with three doses of mRNA COVID-19 vaccines, we observed that a booster dose of the novel sa-mRNA COVID-19 vaccine, ARCT-154, elicited higher immune responses against Wuhan-Hu-1 SARS-CoV-2 that met the prespecified non inferior criteria compared with those elicited by the Comirnaty mRNA vaccine. Responses against the Omicron BA.4/5 variant met all criteria showing they were superior to those elicited by Comirnaty. Furthermore, both ARCT-154 and Comirnaty were well tolerated in this adult population, with no reports of deaths, serious adverse events or medically-attended adverse events causally associated with vaccination.

While heterologous boosting of some COVID-19 vaccines has been shown to improve both the magnitude and breadth of the immune response when compared with homologous boosters [15, 16], this has not been the case for mRNA vaccines, where the response to a homologous mRNA booster is usually superior to the response to a heterologous non-mRNA booster [7–10]. However, the ongoing evolution and emergence of new SARS-CoV-2 variants that may be less susceptible to the immunity induced by current mRNA vaccines will require future immunisation campaigns to consider updated strategies, potentially including annual boosters with different and new vaccines to enhance the breadth of the immune response and minimize vaccine evasion.

The present study was intended to investigate the impact of one such new vaccine, ARCT-154, when administered as a heterologous booster in adults previously fully immunised with three doses of mRNA vaccines. ARCT-154 is a sa-mRNA vaccine, that uses the Venezuela equine encephalitis virus engineered with an RNA replicon coding for the spike (S) glycoprotein of SARS-CoV-2 D614G variant to replace the RNA coding for the viral structural proteins. By self-generating the S protein, ARCT-154 is intended to provide a similar or improved level and extended duration of antigen expression at a lower vaccine load than an equivalent mRNA vaccine, 5 μg vs 30 μg in the present study [11–13].

We have shown that ARCT-154 elicited an immune response against the ancestral SARS-CoV-2 strain, Wuhan-Hu-1, that was higher than the mRNA vaccine, Comirnaty. More importantly, the response induced by ARCT-154 against a recent variant, Omicron BA.4/5, was shown to be superior to that elicited by Comirnaty. This new sa-mRNA platform appears to induce higher heterologous responses, a characteristic which is highly desirable in a period of continued viral evolution. The improved immune response was observed across subgroups, including in men and women and in older individuals (aged 65 years or older) although the low numbers of participants in some groups, e.g., only 19 participants were ≥ 65 years of age, suggest caution should be exercised in the interpretation of these data (see *supplementary tables 4–6*).

This immunogenicity was not accompanied by any observed differences in safety or reactogenicity profile. This finding is consistent with data from the major phase1/2/3/3b study (ARCT-154; NCT05012943), which demonstrated a vaccine safety profile similar to that of the placebo (manuscript in preparation). Both ARCT-154 and Comirnaty were well tolerated with mainly mild to moderate solicited adverse events, similar to those reported in previous studies of mRNA vaccine boosters [15,16]. Most such events were local reactions, principally pain or tenderness at the injection site, which rapidly resolved without further sequelae. There were also reports of transient mild to moderate systemic adverse events, notably malaise and headache, which occurred with similar frequencies with both vaccines. There were no vaccine-related serious adverse events or withdrawals due to adverse events. Safety observations are ongoing and are intended to follow up vaccinees for one year after vaccination. As mRNA COVID-19 vaccines have been associated with cases of myocarditis in young adults [17], we monitored for chest pain and shortness of breath in the 7 days after vaccination as indicators of myocarditis and pericarditis. Six participants, two ARCT-154 and four Comirnaty recipients, reported one or both symptoms. Although one ARCT-154 case and all four Comirnaty cases were considered to be related to the study vaccines, myocarditis and pericarditis were excluded following evaluation by the investigators or a cardiologist.

Our study does have some limitations, notably the already mentioned low numbers of older participants which precludes a meaningful analysis of the response in the elderly and will require a further assessment in this population. Although immunogenicity was assessed as neutralising antibodies against both the original Wuhan-Hu-1 SARS-CoV-2 virus and the Omicron BA.4/5 variant, protective responses against the currently predominating variants (Omicron XBB.1.5-like and Omicron XBB.1.16) need to be assessed and such studies are currently being planned. As already noted, this interim assessment has only monitored safety and immunogenicity up to Day 29, but the study is ongoing to collect safety data and will also assess the durability of the immune response at 3, 6 and 12 months postvaccination.

In conclusion, the novel ARCT-154 sa-mRNA vaccine was well tolerated by healthy adults, and four weeks after vaccination of mRNA-primed adults, one dose elicited a neutralising antibody response against Wuhan-Hu-1 SARS-CoV-2 that was non-inferior to a homologous booster with Comirnaty vaccine. Importantly, the neutralising response against Omicron BA.4/5 variant following ARCT-154 administration was superior to that observed following a booster dose of Comirnaty. The development of mRNA vaccines against COVID-19 has been a success; however, new technologies – such as sa-mRNA – can help to further reduce the burden of disease. The data presented in this report demonstrates the sa-mRNA vaccine technology induces higher antibody titres against key variants, and consequently will potentially enhance the duration of protection.

## Data Availability

All data produced in the present work are contained in the manuscript

## Contributors

YO, YK, MK, JW, BG and YZ all participated in the conception and design of the study and contributed to the writing of the protocol. YO, MK and IO contributed to study oversight. YI performed all statistical elements of study design and analysis of the results. YY was responsible for project administration. TK contributed to funding acquisition. All authors contributed to the interpretation of the results. JW led the writing of the initial drafts of the manuscript on which all authors commented, and all authors agreed with the decision to submit the final manuscript for publication.

## Conflicts of interest

YO, MK, YI, IO, TM and YY are full-time employees and TK is a board member of the study sponsor. YK received fees from Meiji Seika Pharma Co., Ltd., for medical consultation during this study. BFG and YZ are full-time employees of Arcturus Therapeutics, Inc., who developed the vaccine, JLW is an independent consultant working for Arcturus Therapeutics, Inc. Other authors have no conflicts to declare.

## ACKNOWLEDGEMENTS

The authors wish to thank all participants for volunteering, and all study staff and investigators (Takashi Eto, Kenji Takazawa, Hiroaki Kondo, Kenichi Furihata, Hidetoshi Furuie, Osamu Matsuoka, Shinya Mitsui, Yuki Sekiguchi, Shokei Kim-Mitsuyama, Masao Kobayakawa, Toshio Naito) for their skillful application to the study. We are grateful to Keith Veitch (keithveitch communications, Amsterdam, The Netherlands) for editorial assistance in the preparation of the manuscript, and to Jonathan Edelman (Seqirus Inc., USA) and Igor Smolenov (Arcturus Therapeutics, Inc. USA) for critical review.

## Notes

### Clinical Trial

jRCT 2071220080

### Author Declarations

Hakata Clinic Institutional Review Board of Hakata CL gave ethical approval for this work Institutional review board of Shinanozaka clinic of Shinanozaka CL gave ethical approval for this work P-One Clinic, Keikokai Medical Corp Institutional Review Board of Higashi-Shinjuku CL gave ethical approval for this work P-One Clinic, Keikokai Medical Corp Institutional Review Board of P-One CL gave ethical approval for this work Medical Corporation Heishinkai OPHAC Hospital Institutional Review Board of Osaka Pharmacology Clinical Research Hospital gave ethical approval for this work Medical Corporation Heishinkai OPHAC Hospital Institutional Review Board of ToCROM gave ethical approval for this work Kobori Central Clinical Research Ethics Committee of Shin-Sapporo HP gave ethical approval for this work Kobori Central Clinical Research Ethics Committee of LUNA gave ethical approval for this work Kobori Central Clinical Research Ethics Committee of Medimesse Sakurajyuji gave ethical approval for this work Fukushima Medical University Hospital Institutional Review Board of Fukushima Medical HP gave ethical approval for this work Juntendo University Hospital Institutional Review Board of Juntendo HP gave ethical approval for this work

