## Supplementary appendix for "Booster dose of self-amplifying SARS-CoV-2 RNA vaccine vs. mRNA vaccine: a phase 3 comparison of ARCT-154 with Comirnaty^®^"

### Supplementary material


**Table 1:** Study sites and locations page 2

**Table 2:** Severity scale for local reactions page 3

**Table 3:** Severity scale for systemic adverse events page 4

**Table 4:** Immunogenicity comparisons according to age category page 5

**Table 5:** Immunogenicity comparisons according to gender page 6

**Table 6:** Immunogenicity comparisons according to interval since last vaccination page 7

**Table 7:** Duration of local reactions page 8

**Table 8:** Duration of systemic adverse events page 9

**Table 9:** Unsolicited adverse events according to MedDRA page 10

**Supplementary table 1.** Study sites and ethical committees for the study.

| Site No. | Site name | Location | Ethical committee |
|---|---|---|---|
| **1** | Hakata CL | Fukuoka | Hakata Clinic Institutional Review Board |
| **2** | Shinanozaka CL | Tokyo | Institutional review board of Shinanozaka clinic |
| **3** | Higashi-Shinjuku CL | Tokyo | P-One Clinic, Keikokai Medical Corp. IRB |
| **4** | P-One CL | Tokyo | |
| **5** | Osaka Pharmacology Clinical Research Hospital | Osaka | Medical Corporation Heishinkai OPHAC Hospital IRB |
| **6** | ToCROM | Tokyo | |
| **7** | Shin-Sapporo HP | Hokkaido | Kobori Central Clinical Research Ethics Committee |
| **8** | LUNA | Kanagawa | |
| **9** | Medimesse Sakurajyuji | Kumamoto | |
| **10** | Fukushima Medical HP | Fukushima | Fukushima Medical University Hospital Institutional Review Board |
| **11** | Juntendo HP | Tokyo | Juntendo University Hospital Institutional Review Board |

**Supplementary table 2.** Severity scale for local reactions

| **Local Reaction** | **Mild** (Grade 1) | **Moderate** (Grade 2) | **Severe** (Grade 3) | **Potentially life threatening** (Grade 4) |
|---|---|---|---|---|
| Pain | Does not interfere with activity | Repeated use of non-narcotic pain reliever > 24 hours or interferes with activity | Any use of narcotic pain reliever or prevents daily activity | Emergency room (ER) visit or hospitalisation |
| Tenderness | Mild discomfort to touch | Discomfort with movement | Significant discomfort at rest | ER visit or hospitalisation |
| Erythema / Redness | 2·5 – 5·0 cm | 5·1 – 10 cm | > 10 cm | Necrosis or exfoliative dermatitis |
| Induration / Swelling | 2·5 – 5·0 cm and does not interfere with activity | 5·1 – 10 cm or interferes with activity | > 10 cm or prevents daily activity | Necrosis |

**Supplementary table 3.** Severity scale for systemic adverse events

| **Systemic adverse event** | **Mild** (Grade 1) | **Moderate** (Grade 2) | **Severe** (Grade 3) | **Potentially life threatening** (Grade 4) |
|---|---|---|---|---|
| Fever (°C) | 38·0° – 38·4° | 38·5° – 38·9° | 39·0° – 40·0° | > 40·0° |
| Nausea / vomiting | No interference with activity or 1 – 2 episodes/24 hours | Some interference with activity or > 2 episodes/24 hours | Prevents daily activity, requires outpatient IV hydration | ER visit or hospitalisation for hypotensive shock |
| Diarrhoea | 2 – 3 loose stools or < 400 g/24 hr | 4 – 5 stools or 400 – 800 g/24 hr | 6 or more watery stools or > 800 g/24 hr or requires outpatient IV hydration | ER visit or hospitalisation |
| Headache | No interference with activity | Repeated use of non- narcotic pain reliever > 24 hours or some interference with activity | Significant; any use of narcotic pain reliever or prevents daily activity | ER visit or hospitalisation |
| Fatigue | No interference with activity | Some interference with activity | Significant; prevents daily activity | ER visit or hospitalisation |
| Myalgia | No interference with activity | Some interference with activity | Significant; prevents daily activity | ER visit or hospitalisation |
| Dizziness, arthralgia, chills | No interference with activity | Some interference with activity not requiring medical intervention | Prevents daily activity and requires medical intervention | ER visit or hospitalisation |

**Supplementary table 4.**

Geometric mean titres (GMT) of neutralising antibodies and seroresponse rates (SRR) at Day 29 with non-inferiority comparisons after ARCT-154 and Comirnaty (PPS-1) – **according to age category**

| | | **GMT** (95% CI) | | **Seroresponse rate** n / N **%** (95% CI) | |
|---|---|---|---|---|---|
| **Assay** | **Booster vaccine =** | **ARCT-154** | **Comirnaty** | **ARCT-154** | **Comirnaty** |
| **Neutralising antibodies against SARS-CoV-2 Wuhan-Hu-1** | | | | | |
| **< 65 years of age** | | N = 374<br>**5628**<br>(4310, 7349) | N = 366<br>**3953**<br>(3007, 5197) | 244 / 374<br>**65·2%**<br>(60·2, 70·1) | 188 / 366<br>**51·4%**<br>(46·1, 56·6) |
| **GMT ratio or SRR difference** (95% CI) | | **1·42** (1·25, 1·62) | | **13·9%** (6·8, 20·8) | |
| **≥ 65 years of age** | | N = 11<br>**6818**<br>(4055, 11464) | N = 8<br>**3218**<br>(1684, 6149) | 7 / 11<br>**63·6%**<br>(30·8, 89·1) | 5 / 8<br>**62·5%**<br>(24·5, 91·5) |
| **GMT ratio or SRR difference** (95% CI) | | **2·12** (0·92, 4·87) | | **1·1%** (-39·7, 43·3) | |
| **Neutralising antibodies against SARS-CoV-2 Omicron BA.4/5** | | | | | |
| **< 65 years of age** | | N = 374<br>**2556**<br>(1689, 3870) | N = 366<br>**1967**<br>(1286, 3010) | 261 / 374<br>**69·8%**<br>(64·9, 74·4) | 210 / 366<br>**57·4%**<br>(52·1, 62·5) |
| **GMT ratio or SRR difference** (95% CI) | | **1.30** (1·07, 1·58) | | **12.4%** (5·5, 19·2) | |
| **≥ 65 years of age** | | N = 11<br>**2616**<br>(1180, 5801) | N = 8<br>**2053**<br>(761, 5537) | 8 / 11<br>**72·7%**<br>(39·0, 94·0) | 7 / 8<br>**87·5%**<br>(47·3, 99·7) |
| **GMT ratio or SRR difference** (95% CI) | | **1·27** (0·36, 4·56) | | **-14·8%** (-48·9, 26·7) | |

* GMT and GMT ratio calculated using an ANCOVA model.

**Supplementary table 5.**

Geometric mean titres (GMT) of neutralising antibodies and seroresponse rates (SRR) at Day 29 with non-inferiority comparisons after ARCT-154 and Comirnaty (PPS-1) – **striated by Gender**

| | | **GMT** (95% CI) | | **Seroresponse rate** n / N **%** (95% CI) | |
|---|---|---|---|---|---|
| **Assay** | **Booster vaccine =** | **ARCT-154** | **Comirnaty** | **ARCT-154** | **Comirnaty** |
| **Neutralising antibodies against SARS-CoV-2 Wuhan-Hu-1** | | | | | |
| **Male** | | N = 157 **7433** (5003, 11041) | N = 162 **5107** (3426, 7613) | 106 / 157 **67·5%** (59·6, 74·8) | 89 / 162 **54·9%** (46·9, 62·8) |
| **GMT ratio or SRR difference** (95% CI) | | **1·46** (1·20, 1·76) | | **12·6%** (1·9, 23·0) | |
| **Female** | | N = 228 **4481** (3119, 6437) | N = 212 **3138** (2156, 4566) | 145 / 228 **63·6%** (57·0, 69·8) | 104 / 212 **49·1%** (42·1, 56·0) |
| **GMT ratio or SRR difference** (95% CI) | | **1·43** (1·21, 1·69) | | **14·5%** (5·3, 23·6) | |
| **Neutralising antibodies against SARS-CoV-2 Omicron BA.4/5** | | | | | |
| **Male** | | N = 157 **3001** (1592, 5657) | N = 162 **2549** (1345, 4833) | 109 / 157 **69·4%** (61·6, 76·5) | 95 / 162 **58·6%** (50·6, 66·3) |
| **GMT ratio or SRR difference** (95% CI) | | **1·18** (0·86, 1·60) | | **10·8%** (0·2, 21·1) | |
| **Female** | | N = 228 **2214** (1277, 3838) | N = 212 **1572** (889, 2779) | 160 / 228 **70·2%** (63·8, 76·0) | 122 / 212 **57·5%** (50·6, 64·3) |
| **GMT ratio or SRR difference** (95% CI) | | **1·41** (1·09, 1·81) | | **12·6%** (3·7, 21·5) | |

* GMT and GMT ratio calculated using an ANCOVA model.

**Supplementary table 6.**

Geometric mean titres (GMT) of neutralising antibodies and seroresponse rates (SRR) at Day 29 with non-inferiority comparisons after ARCT-154 and Comirnaty (PPS-1) – **striated according to time since last vaccination**

| | | **GMT** (95% CI) | | **Seroresponse rate** n / N **%** (95% CI) | |
|---|---|---|---|---|---|
| **Assay** | **Booster vaccine =** | **ARCT-154** | **Comirnaty** | **ARCT-154** | **Comirnaty** |
| **Neutralising antibodies against SARS-CoV-2 Wuhan-Hu-1** | | | | | |
| **< 5 months since last vaccination** | | N = 8<br>**5500**<br>(2150, 14069) | N = 3<br>**5304**<br>(1144, 24600) | 3 / 8<br>**37·5%**<br>(8·5, 75·5) | 0 / 3<br>**0**<br>(0, 70·8) |
| **GMT ratio or SRR difference** (95% CI) | | **1·04** (0.16, 6.64) | | **37·5%** (-29·3, 70·6) | |
| **≥ 5 months since last vaccination** | | N = 377<br>**5485**<br>(5016, 5998) | N = 371<br>**3801**<br>(3475, 4158) | 248 / 377<br>**65·8%**<br>(60·8, 70·6) | 193 / 371<br>**52·0%**<br>(46·8, 57·2) |
| **GMT ratio or SRR difference** (95% CI) | | **1·44** (1·27, 1·64) | | **13·8%** (6·7, 20·7) | |
| **Neutralising antibodies against SARS-CoV-2 Omicron BA.4/5** | | | | | |
| **< 5 months since last vaccination** | | N = 8<br>**2339**<br>(560, 9776) | N = 3<br>**5146**<br>(498, 53218) | 4 / 8<br>**50·0**<br>(15·7, 84·3) | 1 / 3<br>**33·3**<br>(0·8, 90·6) |
| **GMT ratio or SRR difference** (95% CI) | | **0·45** (0·03, 7·68) | | **16·7%** (-44·4, 63·6) | |
| **≥ 5 months since last vaccination** | | N = 377<br>**2161**<br>(1881, 2483) | N = 371<br>**1635**<br>(1422, 1880) | 265 / 377<br>**70·3%**<br>(65·4, 74·9) | 216 / 371<br>**58·2%**<br>(53·0, 63·3) |
| **GMT ratio or SRR difference** (95% CI) | | **1·32** (1·09, 1·61) | | **12·1%** (5·2, 18·8) | |

* GMT and GMT ratio calculated using an ANCOVA model.

**Supplementary table 7.**

Duration of solicited local reactions in days (Safety Set).

| | | ARCT-154 | Comirnaty |
|---|---|---|---|
| | n | 398 | 395 |
| **Any solicited local reaction** | **Mean ± SD** | **4·7 ± 2·4** | **4·5 ± 1·6** |
| | Median | 4·0 | 4·0 |
| | n | 352 | 358 |
| Injection site pain | **Mean ± SD** | **3·5 ± 1·4** | **3·6 ± 1·4** |
| | Median | 3·0 | 3·0 |
| | n | 388 | 391 |
| Injection site tenderness | **Mean ± SD** | **4·6 ± 2·3** | **4·4 ± 1·6** |
| | Median | 4·0 | 4·0 |
| | n | 59 | 97 |
| Injection site pain swelling | **Mean ± SD** | **3·0 ± 1·2** | **3·4 ± 1·7** |
| | Median | 3·0 | 3·0 |
| | n | 52 | 85 |
| Injection site erythema | **Mean ± SD** | **4·9 ± 2·8** | **3·2 ± 1·7** |
| | Median | 5·0 | 3·0 |
| | n | 52 | 81 |
| Injection site induration | **Mean ± SD** | **3·9 ± 1·8** | **3·7 ± 2·1** |
| | Median | 3·0 | 3·0 |

**Supplementary table 8.**

Duration of solicited systemic adverse events in days

| | | ARCT-154 | Comirnaty |
|---|---|---|---|
| | n events | 276 | 255 |
| **Any solicited systemic AE** | **Mean ± SD** | **3·0 ± 1·4** | **3·0 ± 1·5** |
| | Median | 3·0 | 2·0 |
| | n events | 84 | 76 |
| Pyrexia | **Mean ± SD** | **2·0 ± 0·5** | **1·8 ± 0·6** |
| | Median | 2·0 | 2·0 |
| | n events | 112 | 113 |
| Arthralgia | **Mean ± SD** | **2·6 ± 1·2** | **2·5 ± 1·2** |
| | Median | 2·0 | 2·0 |
| | n events | 126 | 103 |
| Chills | **Mean ± SD** | **2·4 ± 0·9** | **2·2 ± 0·5** |
| | Median | 2·0 | 2·0 |
| | n events | 28 | 17 |
| Diarrhoea | **Mean ± SD** | **2·5 ± 1·0** | **3·0 ± 2·2** |
| | Median | 2·0 | 2·0 |
| | n events | 25 | 13 |
| Dizziness | **Mean ± SD** | **2·8 ± 1·2** | **2·5 ± 1·2** |
| | Median | 2·0 | 2·0 |
| | n events | 165 | 125 |
| Headache | **Mean ± SD** | **2·7 ± 1·1** | **2·8 ± 1·5** |
| | Median | 2·0 | 2·0 |
| | n events | 188 | 176 |
| Malaise | **Mean ± SD** | **2·9 ± 1·4** | **2·7 ± 1·3** |
| | Median | 2·0 | 2·0 |
| | n events | 21 | 16 |
| Nausea | **Mean ± SD** | **2·4 ± 0·9** | **2·7 ± 2·0** |
| | Median | 2·0 | 2·0 |
| | n events | 2 | 2 |
| Vomiting | **Mean ± SD** | **2·0 ± 0** | **2·0 ± 0** |
| | Median | 2·0 | 2·0 |
| | n events | 123 | 100 |
| Myalgia | **Mean ± SD** | **3·9 ± 1·8** | **3·7 ± 2·1** |
| | Median | 2·0 | 3·0 |

**Supplementary table 9.**

Rates of unsolicited adverse events according to MedDRA classifications in the two study groups from Day 1 to Day 29 (FAS)

| | **ARCT-154** | **Comirnaty** |
|---|---|---|
| N = | 420 | 408 |
| **Any unsolicited adverse event,** n participants (%) | 81 (19·3) | 111 (27·2) |
| Blood and lymphatic disorders | 2 (0·5) | 7 (1·7) |
| Cardiac disorders | 3 (0·7) | 0 |
| Eye disorders | 2 (0·5) | 2 (0·5) |
| Gastrointestinal disorders | 11 (2·6) | 13 (3·2) |
| General disorders and administration site conditions | 28 (6·7) | 47 (11·.5) |
| Hepatobiliary disorders | 1 (0·2) | 0 |
| Immune system disorders | 1 (0·2) | 1 (0·2) |
| Infections and infestations | 7 (1·7) | 14 (3·4) |
| Injury, poisoning and procedural complications | 2 (0·5) | 3 (0·7) |
| Investigations | 5 (1·2) | 6 (1·5) |
| Metabolism and nutrition disorders | 1 (0·2) | 0 |
| Musculoskeletal and connective tissue disorders | 10 (2·4) | 15 (3·7) |
| Nervous system disorders | 11 (2·6) | 9 (2·2) |
| Psychiatric disorders | 0 | 1 (0·2) |
| Renal and urinary disorders | 0 | 1 (0·2) |
| Reproductive system and breast disorders | 1 (0·2) | 1 (0·2) |
| Respiratory, thoracic and mediastinal disorders | 8 (1·9) | 8 (2·0) |
| Skin and subcutaneous tissue disorders | 6 (1·4) | 5 (1·2) |
| Vascular disorders | 1 (0·2) | 3 (0·7) |
